# Genetic landscape of Parkinson’s disease in the Personalized Parkinson Project cohort

**DOI:** 10.64898/2026.03.16.26348127

**Authors:** Theresa Lüth, Christine Klein, Rick C. Helmich, Norbert Brüggemann, Sana Hrir, H. Bea Kuiperij, Vyron Gorgogietas, Sara Beatriz Gomes Fernandes, Jannik Prasuhn, Patrick May, Tiago F. Outeiro, Kenan Steidel, Zied Landoulsi, Teresa Kleinz, Susen Schaake, Christoph Much, Rejko Krüger, Marcel M. Verbeek, Bastiaan R. Bloem, Bart P. van de Warrenburg, Joanne Trinh

## Abstract

**Background:** Parkinson’s disease (PD) is a multifactorial neurodegenerative disorder shaped by, amongst others, high-impact variants and common polygenic factors. The Personalized Parkinson Project (PPP) offers deep phenotyping and longitudinal follow-up of Dutch people with PD. Here, we characterize the genetic landscape and its interaction with lifestyle factors within PPP.

**Methods:** We utilized three complementary approaches in N=507 persons with PD: 1) short-read PD gene panel sequencing of eight PD genes, 2) genome-wide genotyping array, and 3) targeted long-read sequencing of the *GBA1* gene. Additionally, we calculated the mitochondrial-function polygenic score (MGS). Associations between genetic factors, smoking status, and age at onset (AAO) were assessed using non-parametric tests, correlation analyses, and multiple regression models.

**Results:** Genetic screening of the participants revealed N=79 *GBA1* (∼15%), N=3 *LRRK2*, N=1 *CHCHD2*, N=1 *SNCA* variant carrier, and N=9 heterozygous *PRKN/PINK1* variants. We also observed an interaction between MGS and smoking: MGS was associated with earlier AAO in non-smokers in persons with iPD (N=414, β=-1.87, p=0.038).

**Conclusion:** Our findings corroborate previously reported frequencies of variants in PD-associated genes in European populations, and suggest a potential association between smoking and a mitochondrial dysfunction signature in PD. Thus, even in persons without rare variants (iPD subgroup), complex genetic contributions remained relevant. Our study supports future downstream stratification and personalized medicine approaches with high-impact variants and polygenic risk scores.

## Introduction

Parkinson’s Disease (PD) is a progressive and multifactorial neurodegenerative disorder with a complex etiology. It is presently the fastest-growing neurodegenerative disease as of today, with almost 12 million patients worldwide^1^. Reliable multi-time-point longitudinal datasets that capture both the epidemiology and genetic underpinnings of PD are critically lacking. Comprehensive, deep characterization of current PD-associated genes in rigorously phenotyped cohorts is indispensable for advancing gene-targeted clinical trials, enabling precision medicine, and facilitating molecular risk profiling.

Recent studies have quantified the genetic contribution to PD across populations, showing that up to 14.8% of cases are attributable to *GBA1* variants, representing the largest proportion^2, 3^. These findings highlight that single impactful variants remain an important component of PD genetics. However, PD susceptibility is not determined solely by single variants. Common genetic polygenic factors also shape PD risk^4, 5^. Mitochondrial dysfunction is a key player in PD pathogenesis and has been observed in monogenic as well as idiopathic PD (iPD) forms. Indeed, common variants in nuclear-encoded mitochondrial genes contribute modestly but cumulatively to PD-risk in a polygenic fashion; in detail, the increase of the mitochondrial polygenic score (MGS) by one standard deviation was associated with a 21% higher odds of PD affection status in the European ancestry group^6, 7^. Additionally, related mitochondrial impairments were functionally validated in patient-derived neuronal models^8^. Furthermore, environmental or lifestyle factors and their interactions with genetic variants impact iPD and monogenic forms as well as disease onset and progression^9, 10, 6, 11-13^. Some gene-environment interactions may be mediated through mitochondrial dysfunction. For example, genetically determined deficits in mitochondrial function may increase vulnerability to neurotoxic effects after exposure to pesticides that also target mitochondrial pathways. Our group demonstrated that the MGS was associated with earlier age at onset (AAO) among non-smokers or persons who regularly consume caffeinated soda, in people with iPD or *LRRK2*-related PD, respectively ^6^. Additionally, we reported an additive association between aspirin use and a PD-specific polygenic score ^10^.

Thus, it is essential to investigate variants associated with monogenic PD forms, as well as the contribution of complex polygenic mitochondrial risk, within deeply phenotyped cohorts. The Personalized Parkinson Project (PPP) offers an exceptional resource due to its longitudinal, multidimensional characterization of Dutch people with PD, spanning clinical, biological, environmental, and lifestyle assessments^14^. Investigating both high-impact variants and polygenic contributions within the PPP will therefore provide valuable insights into PD etiology in this well-characterized cohort.

## Methods

### The Personalized Parkinson Project study cohort

DNA from N = 507 participants in the PPP cohort was included in this study to assess PD-relevant genetic variants, all of whom had PD (**Table 1**). The mean age at symptom onset (*AAO*) was 56.2 years (±10.2), and the mean age at initial examination (AAE) was 61.7 years (±9.2). N = 302 participants were men (59.4%). The PPP study includes deep phenotyping data on PD, including motor and non-motor outcomes, biomarkers, and environmental and lifestyle factors, collected longitudinally over a two-year follow-up period^14^. As described previously, participants were eligible if they had a clinical diagnosis of Parkinson’s disease within the past 5 years and were ≥18 years old. Comorbidities were not an exclusion criterion unless they substantially interfered with the assessment of Parkinsonian disability. To ensure a representative cohort, participants were recruited using a stratified inclusion strategy based on sex, age, and disease duration^14^. The study was conducted in conformance with the Good Clinical Practice ICH E6 guideline, and in compliance with the Ethical Principles for Medical Research Involving Human Subjects, as defined in the Declaration of Helsinki (version amended in October 2013), the Dutch Personal Data Protection Act, and the European General Data Protection Regulation. The Commissie Mensgebonden Onderzoek Region Arnhem-Nijmegen (reference number 2016–2934; NL59694.091.17) approved the study protocol and communication materials. Written informed consent was obtained before subjecting a participant to any study procedure.

**Table 1.**
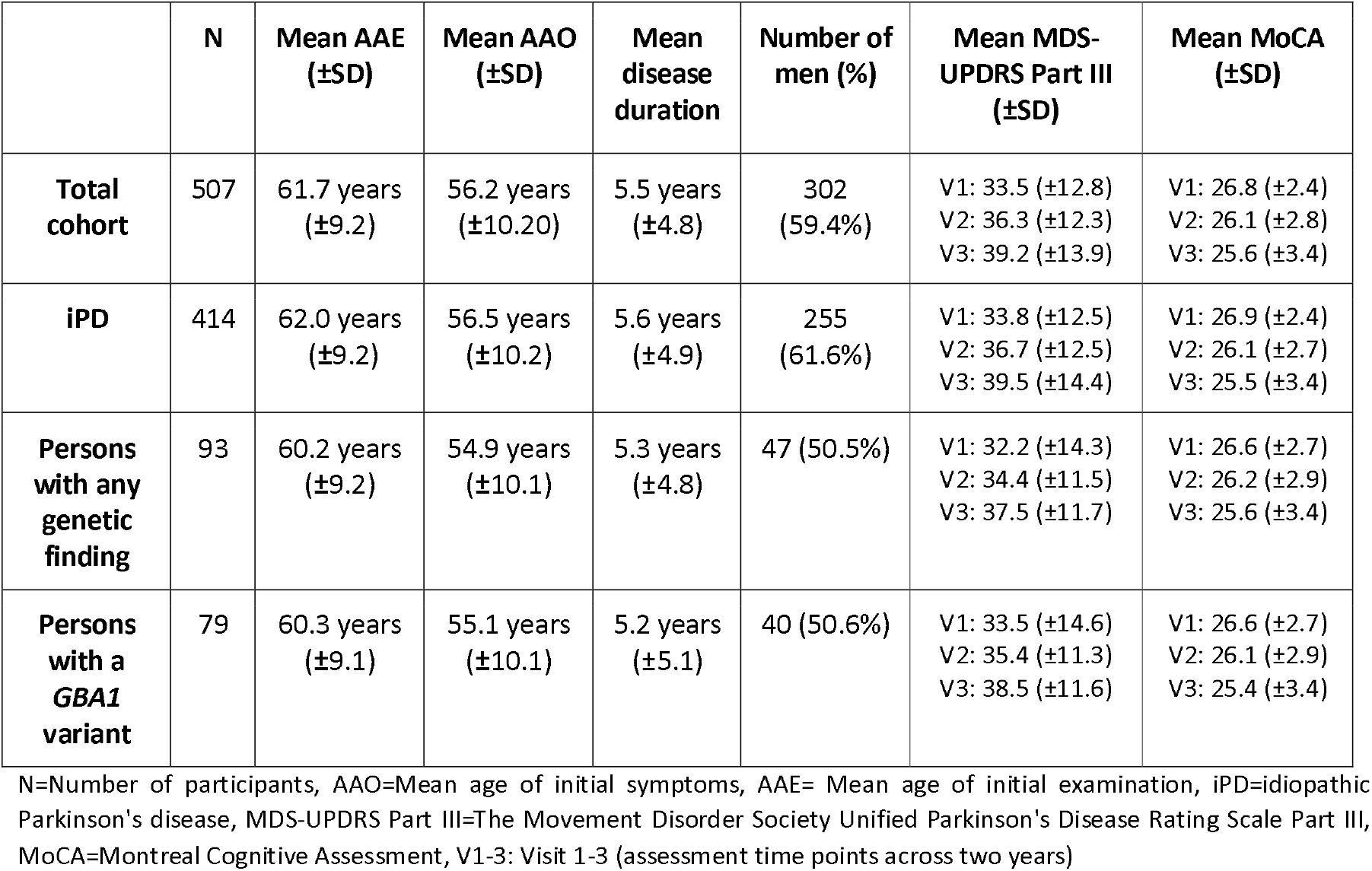
Demographic overview of PPP participants who have been screened for PD-relevant genetic variants.

| | N | Mean AAE<br>(±SD) | Mean AAO<br>(±SD) | Mean<br>disease<br>duration | Number of<br>men (%) | Mean MDS-UPDRS Part III<br>(±SD) | Mean MoCA<br>(±SD) |
| --- | --- | --- | --- | --- | --- | --- | --- |
| **Total cohort** | 507 | 61.7 years<br>(±9.2) | 56.2 years<br>(±10.20) | 5.5 years<br>(±4.8) | 302<br>(59.4%) | V1: 33.5 (±12.8)<br>V2: 36.3 (±12.3)<br>V3: 39.2 (±13.9) | V1: 26.8 (±2.4)<br>V2: 26.1 (±2.8)<br>V3: 25.6 (±3.4) |
| **iPD** | 414 | 62.0 years<br>(±9.2) | 56.5 years<br>(±10.2) | 5.6 years<br>(±4.9) | 255<br>(61.6%) | V1: 33.8 (±12.5)<br>V2: 36.7 (±12.5)<br>V3: 39.5 (±14.4) | V1: 26.9 (±2.4)<br>V2: 26.1 (±2.7)<br>V3: 25.5 (±3.4) |
| **Persons with any genetic finding** | 93 | 60.2 years<br>(±9.2) | 54.9 years<br>(±10.1) | 5.3 years<br>(±4.8) | 47 (50.5%) | V1: 32.2 (±14.3)<br>V2: 34.4 (±11.5)<br>V3: 37.5 (±11.7) | V1: 26.6 (±2.7)<br>V2: 26.2 (±2.9)<br>V3: 25.6 (±3.4) |
| **Persons with a *GBA1* variant** | 79 | 60.3 years<br>(±9.1) | 55.1 years<br>(±10.1) | 5.2 years<br>(±5.1) | 40 (50.6%) | V1: 33.5 (±14.6)<br>V2: 35.4 (±11.3)<br>V3: 38.5 (±11.6) | V1: 26.6 (±2.7)<br>V2: 26.1 (±2.9)<br>V3: 25.4 (±3.4) |

N=Number of participants, AAO=Mean age of initial symptoms, AAE= Mean age of initial examination, iPD=idiopathic Parkinson's disease, MDS-UPDRS Part III=The Movement Disorder Society Unified Parkinson's Disease Rating Scale Part III, MoCA=Montreal Cognitive Assessment, V1-3: Visit 1-3 (assessment time points across two years)

### Detection of PD-relevant genetic findings

We integrated the results from three genetic assessment approaches: 1) targeted PD gene panel, 2) Infinium Global Screening Array (*GSA*) genotyping array with custom content for neurodegenerative diseases and 3) targeted *GBA1* long-read sequencing. To ensure the validity of genetic findings, we required the detection with at least two of the methods, and if that was not possible, we performed Sanger sequencing to confirm the variant as previously described^15^.

### Short-read PD gene panel sequencing

The DNA from three individual persons was placed together into one barcoded pool, and each pool underwent sequencing using a next-generation gene panel at CENTOGENE GmbH. The panel targets 50 genes related to neurological disorders, and eight genes were included in the analysis (i.e., *LRRK2, GBA1, PRKN, PINK1, DJ1, SNCA, VPS35* and *CHCHD2*)^3^ as they are defined as relevant for PD according to the recommendations of the International Parkinson and Movement Disorder Society task force^16^. Variant calling was performed with *Mutec*^17^, and the variants were annotated with GnomAD, RefSeq and pathogenicity prediction tools, including MutTaster, ClinVar, PhyloP and Sift. The variants were filtered based on the annotation items with the following criteria: 1) rare variant frequency in the general population, GnomAD minor allele frequency <0.05; 2) Combined Annotation Dependent Depletion (CADD) score >10^18^; 3) coding changes or splice variants; 4) predicted pathogenicity in two or more additional tools or databases (MutTaster, ClinVar, PhyloP or Sift) ^19-22^.

### Genotyping with the Infinium Global Screening Array

Array-based genotyping was performed on all individual DNA samples, utilizing the Infinium GSA with custom content for neurodegenerative-related variants, assessing ∼700,000 variants. The genotyping data obtained from the GSA array were stored in PLINK format^23^ and imputed using the Michigan Imputation Server (v2, Reference Panel: HRC r1.1, Rsq filter: 0.3) ^24^. Next, principal component analysis was performed, and PC1-2 was clustered with the 1000 Genomes superpopulations^25^ (**Supplementary Figure 1**).

The mitochondrial-function-specific polygenic score (MGS) was calculated for each participant based on the QC-filtered, imputed genotyping data, as previously described^6, 7^. The applied MGS consists of ∼15,000 SNVs located within nuclear-encoded genes associated with mitochondrial function. The single-nucleotide variant (SNV) and the corresponding weight were processed with the *PLINK* score function to obtain the cumulative MGS for each person. All necessary commands, the list of SNVs, and corresponding weights can be found here: https://github.com/LuethTheresa/MitochondrialPolygenicScoreAndAgeAtOnset.

### Long-read sequencing of the *GBA1* gene

Lastly, we performed targeted Oxford Nanopore Technologies (ONT) long-read sequencing of the *GBA1* gene. The long-read assessment of the *GBA1* gene was performed as previously described^26^. In brief, a ∼8.9kb long-range PCR amplicon was generated spanning the entire *GBA1* gene. The library was prepared with the Ligation Sequencing Kit V14 and the Native Barcoding Kit 96 V14. Thus, we barcoded N=96 samples per FLO-PRO114M flow cell.

### Multiplex Ligation-dependent Probe Amplification

Persons in whom we detected heterozygous *PRKN* or *PINK1* variants were further investigated for copy number variations (CNVs) in that gene. CNVs in *PRKN* and *PINK1* were assessed with multiplex ligation-dependent probe amplification (MLPA; MRC Holland, P051 probe mix, including the genes *PRKN, PINK1* and *SNCA*) as described previously^15^.

### Statistical analysis

Statistical analyses were performed with R v4.3.1^27^. Pairwise comparisons were assessed with the non-parametric Wilcoxon-Mann-Whitney test. Associations between MGS and AAO were visualized using Spearman correlation and assessed with multiple linear regression models, adjusting for relevant covariates (i.e., biological sex, PC1-2, and AAE). To assess the longitudinal association of *GBA1* variants with motor and cognitive outcomes, we applied linear mixed-effects models, as described previously^9^. Longitudinal outcomes included motor severity, assessed using the MDS-UPDRS Part III (Movement Disorder Society–Unified Parkinson’s Disease Rating Scale, Part III: Motor Examination), and cognitive performance, assessed using the MoCA (Montreal Cognitive Assessment, a screening tool for global cognitive function). Briefly, MDS-UPDRS Part III scores and MoCA scores were modeled over time with the personal ID as a random effect, and AAO, sex, and principal components 1-2 as fixed effects. Continuous covariates were transformed when appropriate to meet model assumptions.

## Results

### Genetic ancestry and the spectrum of variants within PD genes

Analysis of the first two principal components showed that most PPP participants clustered with the European 1000 Genomes ancestry group, confirming the predominantly European ancestry of the cohort (**Supplementary Figure 1**). Ten samples clustered outside the European ancestry group, consistent with their self-reported Asian, African, or admixed ancestry. We detected variants within PD genes, including *GBA1*, in N=93 out of 507 people with PD from the PPP cohort (18%, **Figure 1**). Out of the 93 variant carriers, one *GBA1* p.Glu365Lys carrier was of Asian-admixed origin.

**Figure 1.**
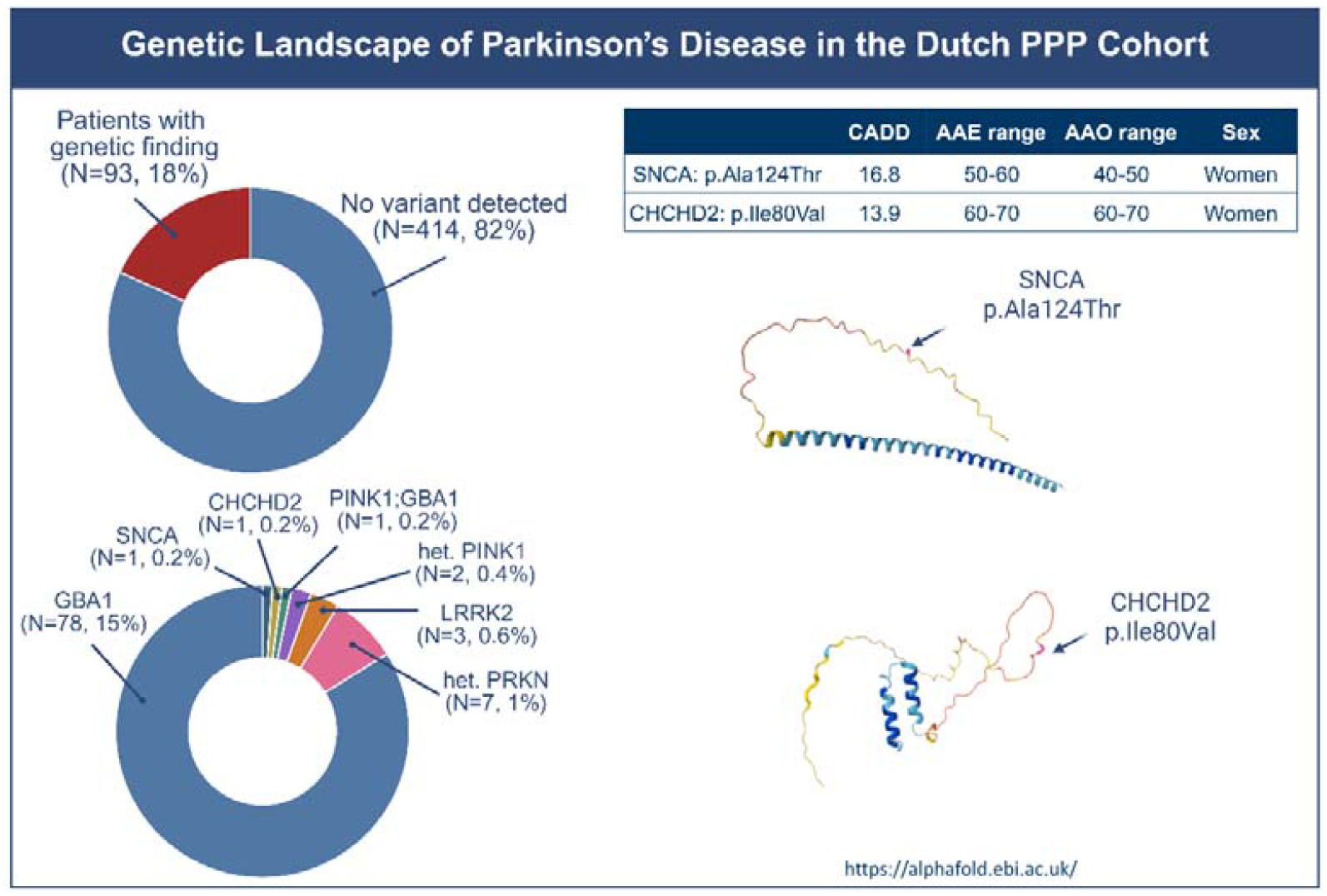
Overview of the frequency and landscape of genetic findings in the Dutch PPP cohort. The figure summarizes the number and percentage of individuals across genetic subgroups. The upper pie chart displays the percentage across all persons with PD in PPP, and the lower chart shows the is an enlargement of the red part with per-gene percentages. It also includes basic demographic information and details on *CHCHD2* and *SNCA* variants, including their CADD scores (SNAC p.Ala124Thr CADD: 16.8, *CHCHD2* p.Ile80Val CADD: 13.9) and protein positions. AAO=Age at onset, AAE=Age at examination.

Among genes associated with autosomal-dominant PD (i.e., *LRRK2, SNCA*, and *CHCHD2*), the LRRK2 p.Gly2019Ser variant was the most common, detected in three persons. In addition, we found one heterozygous rare missense variant in *SNCA* (i.e., p.Ala124Thr, CADD score=16.8) and one rare missense variant in *CHCHD2* (p.Ile80Val, CADD score=13.9; **Table 2**). The *SNCA* and *CHCHD2* missense variant carriers were of European origin. The person carrying the *SNCA* variant did not report a family history of PD, while the person carrying the *CHCHD2* variant did report a family history of PD.

**Table 2.** Overview of detected rare variants in PD-relevant genes and their respective genotypes and frequency in the PPP cohort (N=507 assessed persons with PD).

| Gene | Detected variant & genotype | Number of variant carriers |
| --- | --- | --- |
| ***LRRK2*** | HET:NM_198578.3:p.Gly2019Ser | 3 |
| ***SNCA*** | HET:NM_000345.3:p.Ala124Thr | 1 |
| ***CHCHD2*** | HET:NM_016139.3:p.Ile80Val | 1 |
| ***PRKN*** | HET:NM_004562.2:p.Arg402Cys | 3 |
|  | HET:NM_004562.2:p.Gln34fs | 2 |
|  | HET:NM_004562.2:3_prime_UTR_variant_c.*106G>A | 1 |
|  | HET:NM_004562.2:5_prime_UTR_variant_c.*107G>A | 1 |
| ***PINK1*** | HET:NM_032409.2:p.Ala169Pro | 1 |
|  | HET:NM_032409.2:p.Leu544Pro | 1 |
| ***PINK1;GBA1*** | HET:NM_032409.2:p.Gly411Ser;<br>HET:NM_000157.3:p.Asp492Leu | 1 |

HET=Variant genotype is heterozygous

We did not detect homozygous variants in genes associated with autosomal recessive PD (i.e., *PRKN, PINK1*, and *DJ1*). We found four rare heterozygous *PRKN* variants and three rare heterozygous *PINK1* variants (**Table 2**). All persons with heterozygous *PRKN* or *PINK1* variants (N=10) were assessed using MLPA, but no second copy-number variant was identified.

We next focused on *GBA1*, which represented the largest group (N=79) of genetic variants in the PPP cohort (**Table 3**). Variants within *GBA1* are a significant risk factor for PD, but genetic assessments of this gene locus are complicated by the nearby pseudogene *GBAP1*. The longer read length of Nanopore sequencing and higher mapping accuracy help overcome this issue^26^. The most common *GBA1* variant in the cohort was the known risk variant p.Glu365Lys, detected in 48 persons, almost exclusively in a heterozygous state. Only one person carried the variant in a homozygous state. In total, we identified 15 distinct *GBA1* variants, of which 14 were single-nucleotide variants (SNVs) or small indels. However, we also detected one structural variant (SV) present in two persons (i.e., c.1265_1319del: p.Leu422Profs*4), which is a ∼50bp exonic deletion located in exon 9 of the *GBA1* gene (**Table 3**). That particular SV was only discovered by ONT long-read sequencing and was missed by two other genetic screening approaches (**Supplementary Figure 2**).

**Table 3.** Genotypes and corresponding variant annotations for *GBA1* (transcript: NM_000157.3) observed in the PPP cohort (N=507 assessed persons with PD). Each row represents a unique genotype, including variant annotations from the *GBA1*-PD Browser and MDSGene.

| Genotype | Number of variant carriers | Variant 1 | GBA1-PD browser | MDSGene | Variant 2 | GBA1-PD browser | MDSGene |
| --- | --- | --- | --- | --- | --- | --- | --- |
| HET:p.Glu365Lys | 40 | p.Glu365Lys | Risk | Risk | — | — | — |
| HET:p.Thr408Met | 12 | p.Thr408Met | Risk | Risk | — | — | — |
| HET:p.Asn409Ser | 5 | p.Asn409Ser | Mild | Likely Pathogenic | — | — | — |
| HET:p.Glu365Lys;<br>HET:p.Asp179His | 4 | p.Glu365Lys | Risk | Risk | p.Asp179His | Mild | Likely Pathogenic |
| HET:p.Leu363Pro | 2 | p.Leu363Pro | Unknown | Likely Pathogenic | — | — | — |
| HET:p.Leu483Pro;<br>HET:p.Thr408Met | 2 | p.Leu483Pro | Severe | Pathogenic | p.Thr408Met | Risk | Risk |
| SV:c.1265_1319del:<br>p.Leu422Profs*4 | 2 | p.L422Pfs*4 | Severe | Pathogenic | — | — | — |
| HET:p.Arg502Pro | 1 | p.Arg502Pro | Unknown | Likely Pathogenic | — | — | — |
| HET:p.His350Asn | 1 | p.His350Asn | Unknown | Likely Pathogenic | — | — | — |
| HET:p.Ser527Thr | 1 | p.Ser527Thr | Unknown | VUS | — | — | — |
| HET:p.Thr408Met;<br>HET:p.Glu365Lys | 1 | p.Thr408Met | Risk | Risk | p.Glu365Lys | Risk | Risk |
| HET:p.Val499Met | 1 | p.Val499Met | Unknown | Likely Pathogenic | — | — | — |
| HOM:p.Glu365Lys | 1 | p.Glu365Lys | Risk | Risk | — | — | — |
| HET:p.Glu365Lys;<br>HET:p.Leu483Pro | 1 | p.Glu365Lys | Risk | Risk | p.Leu483Pro | Severe | Pathogenic |
| HET:p.Leu29fs | 1 | p.Leu29fs | Severe | Pathogenic | — | — | — |
| HET:p.Leu483Pro | 1 | p.Leu483Pro | Severe | Pathogenic | — | — | — |
| HET:p.Arg159Trp | 1 | p.Arg159Trp | Severe | Pathogenic | — | — | — |
| HET:p.Trp223Arg;<br>HET:p.Glu365Lys | 1 | p.Trp223Arg | Severe | Pathogenic | p.Glu365Lys | Risk | Risk |
| HET:p.Asp492Leu | 1 | p.Asp492Leu | Unknown | VUS | - | - | - |

HET=Variant genotype is heterozygous, HOM=Variant genotype is homozygous, VUS=Variant of uncertain significance
GBA1-PD Browser: https://ngdc.cncb.ac.cn/databasecommons/database/id/8378
MDSGene: https://www.mdsgene.org/genes/PARK-GBA1

In the remaining individuals (N=414), no genetic findings were detected; these individuals are hereafter referred to as idiopathic PD (iPD).

### Evaluation of pathogenicity for *SNCA* and *CHCHD2* missense variants

Given the uncertain pathogenicity of the *SNCA* p.Ala124Thr and *CHCHD2* p.Ile80Val variants, we integrated external genomic datasets and a literature review. We screened the Accelerating Medicines Partnership PD (AMP-PD) whole-genome data set^28^. The *SNCA* p.Ala124Thr variant was absent from the AMP-PD dataset, consistent with the rarity in population-scale sequencing data (gnomAD allele frequency=1.4×10^-5^). A different base change at the same genomic position (HG38: chr4:89729214, HG19: chr4:90650365 C>A instead of C>T), leading to a p.Ala124Ser missense variant, was found in a healthy control in AMP-PD, who was in their 60’s. The *CHCHD2* p.Ile80Val variant was identified twice in the AMP-PD dataset, once in a patient with PD (AAO in their 70’s) and once in a healthy control.

In addition, we conducted literature reviews to evaluate existing evidence for variants in these genes among people with PD (see **Supplementary Texts 1-2** and **Supplementary Table 1**). Given that the association between *CHCHD2* and PD is relatively recent and remains under debate, we conducted a comprehensive literature review on variants of the entire gene. In contrast, because *SNCA* is a well-established genetic determinant of PD, we restricted our review to variants affecting amino acid residue 124.

No reports of the *SNCA* p.Ala124Thr variant were identified in the literature, indicating a lack of prior evidence for disease association. Instead, only a single report of the related p.Ala124Ser variant was found in a Finnish patient with vascular dementia and schizophrenia, with no established pathogenicity or clear link to PD^29^. To further confirm this finding, we also screened the MDSGene database, which contained no entries corresponding to p.Ala124Thr, suggesting that our study may be the first to report it. We performed a literature review on PD-associated variants in *CHCHD2* and included N=29 publications, of which N=13 reported variants associated with PD and N=2 potential risk factors (**Supplementary Table 1**). These 15 publications included one study reporting the p.Ile80Val in an early-onset PD patient of European ancestry^30^. Combined genetic, population, and literature-based evidence did not permit definitive classification of either missense variant as pathogenic or benign.

### Genotype-phenotype associations in the PPP cohort

Comparing the N=93 persons with any genetic finding with the N=414 persons with iPD, persons with a genetic finding had an approximately 1.5 years earlier AAO (mean AAO (±SD)=54.9 years (±10.1)) compared to persons with iPD (mean AAO (±SD)=56.5 years (±10.2)), but this did not reach statistical significance (Mann-Whitney U-test p-value=0.179). The genetic PD group had a lower percentage of men (persons with a genetic finding: 50.5%, iPD: 61.6%; **Table 1**). The three persons carrying the LRRK2 p.Gly2019Ser variant were of European origin; their AAO ranged from ∼50 to ∼79 years, and the disease duration ranged from 2 to 7 years (**Supplementary Table 2**). The two persons carrying a rare missense variant in *CHCHD2* or *SNCA* were both women with AAOs in their 60’s and 40’s, respectively (**Supplementary Table 2**). None of the *SNCA* or *CHCHD2* variant carriers reported loss or change of sense of taste or smell, but the *CHCHD2* variant carrier reported RBD (baseline REM Sleep Behavior Disorder Screening questionnaire=10).

*GBA1* variant carriers presented the largest group of persons with a genetic finding in the PPP cohort. The mean AAO was approximately one year earlier (mean AAO (±SD)=55.1 years (±10.1)) compared to the mean AAO of people with iPD. We utilized two existing classification schemes to stratify *GBA1* variants: the *GBA1*-variant browser (https://ngdc.cncb.ac.cn/databasecommons/database/id/8378) and the MDS-Gene database (https://www.mdsgene.org/genes/PARK-GBA1), summarized in **Table 3**. To assess differences in PD symptoms or signs across distinct groups of *GBA1* variants, we utilized the MDS-Gene classification (pathogenic, likely pathogenic, risk variant, or VUS) in the following.

Among the N=79 persons carrying a *GBA1* variant, N=9 carried a pathogenic variant, N=14 a likely pathogenic variant, N=54 a risk variant and N=2 a variant of unknown significance (VUS) (**Table 3**). The persons carrying a VUS had the highest AAO among the *GBA1* variant carriers (mean AAO (±SD)=67.5 years (±3.54)), followed by persons with a risk variant (mean AAO (±SD)=56.0 years (±9.82)), persons with a likely pathogenic variant (mean AAO (±SD)=53.5 years (±9.67)), and persons with a pathogenic variant (mean AAO (±SD)=49.1 years (±10.5)). Adjusting for sex and PC1-2, persons with a pathogenic variant had a younger AAO compared to persons with iPD (β=-7.24, SE=3.43, p=0.034).

We observed increases in motor signs (higher MDS-UPDRS part III) and mild cognitive impairment (lower MoCA) across the three assessment time points in all participants with PD (**Table 1**, **Supplementary Figure 3**). We explored the association of *GBA1* variants with motor symptoms and cognitive impairment over time, as assessed using the MDS-UPDRS Part III or the MoCA. Neither risk, likely pathogenic, nor pathogenic variants were associated with changes in motor symptom severity across time (**Supplementary Figure 3A, Supplementary Table 3**). There was a subtle trend for more severe cognitive impairment in persons with pathogenic *GBA1* variants (N=7) compared to iPD patients across three assessment time points (β=-0.06, SE=0.03, p=0.090, **Supplementary Figure 3B, Supplementary Table 3**). The groups appear visually separated (**Supplementary Figure 3B**), but in the mixed linear model, cognitive performance did not differ between carriers of pathogenic/likely pathogenic *GBA1* variants and carriers of *GBA1* risk variants (β = -0.03, SE = 0.03, p = 0.294).

### Association between mitochondrial-function polygenic score, smoking and AAO

Lastly, we investigated complex genetic factors and interactions with lifestyle in persons without known mutations and risk variants (i.e., iPD) in an exploratory manner. We calculated an MGS score for the PPP participants with iPD (N=414), using imputed genotyping data to explore the cumulative impact of more common variants in genes associated with mitochondrial function. In a correlation analysis, we observed no correlation between an MGS and an AAO when assessing all people with iPD (r=-0.05, p=0.325; **Supplementary Figure 4**).

Mitochondria represent a central interface through which environmental and lifestyle factors shape cellular function. Comparing the AAO of persons with iPD from the PPP cohort stratified by smoking status (if participants ever smoked in their lifetime) revealed that persons who smoked had a six-year later AAO (N=238, median AAO (IQR)=60.0 (53.0-65.0)) compared to persons who did not smoke (N=158, median AAO (IQR)=54.0 (48.0-62.0), p=2.4 × 10^-5^, **Figure 2A**). The association between smoking and a later AAO remained when adjusting for sex and general genetic background (β=4.48, SE=1.01, p=1.1 × 10^-5^, **Table 4**). Additionally, we observed an interaction between MGS and smoking status on AAO in people with iPD (β=2.29, SE=1.02, p=0.025, **Table 4**).

**Figure 2.**
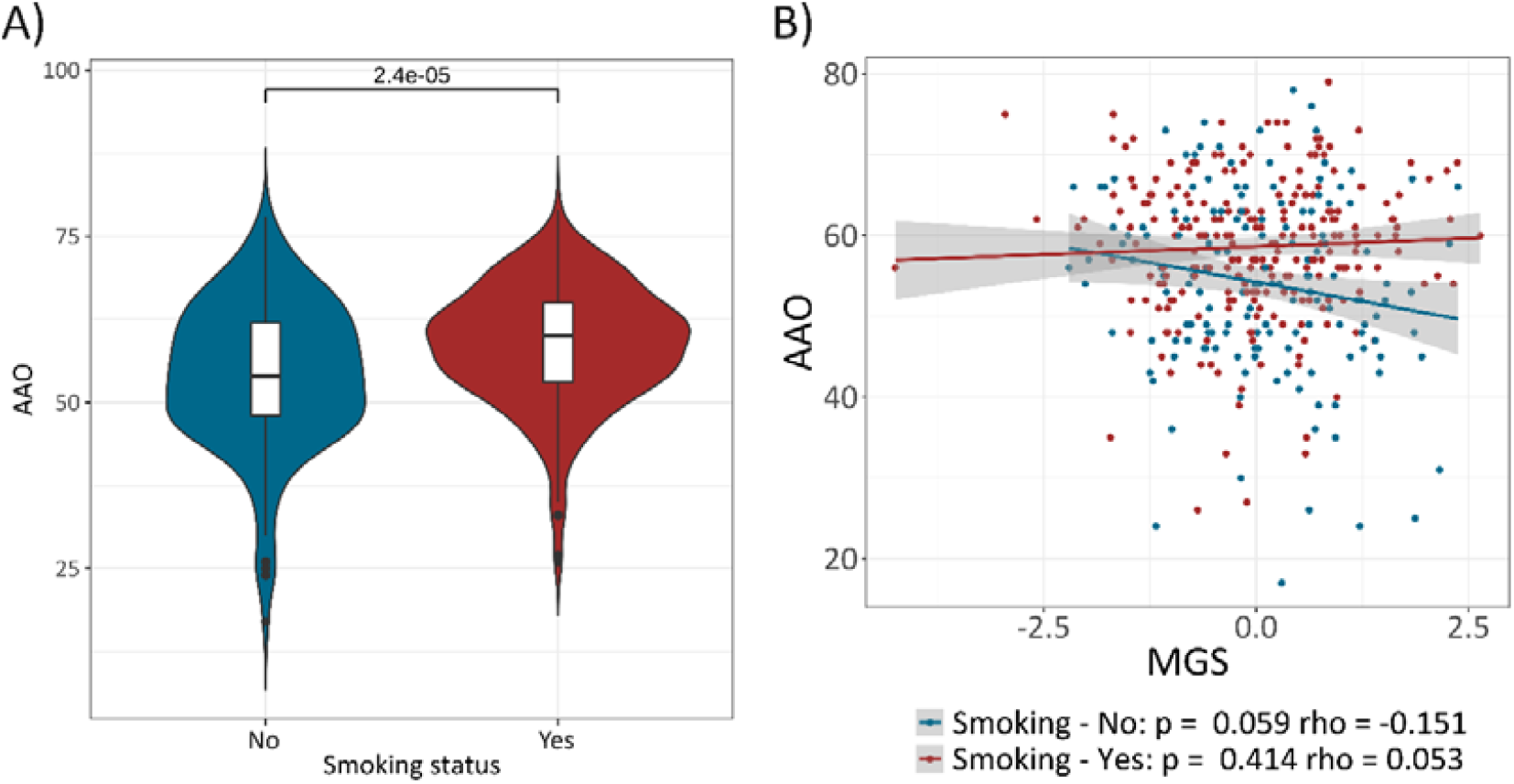
Relationship between the smoking status, mitochondrial polygenic score (MGS) and age of idiopathic PD onset (AAO) and their interaction. (A) The violin plot shows the difference in iPD AAO between persons who smoke and those who do not smoke. The dashed lines indicate the median and interquartile range, and the Mann-Whitney U-Test p-value is displayed. (B) The correlation plots show the association between MGS and AAO in persons with idiopathic Parkinson’s disease (iPD) stratified by smoking status (B). rho=Spearman’s rank correlation coefficient, p=Spearman’s exploratory P-value.

**Table 4.** Relationship between the smoking status, mitochondrial polygenic score (MGS) and age of idiopathic PD onset (AAO) and their interaction.

|  | Estimate | Std. Error | p-value |
| --- | --- | --- | --- |
| **Association between smoking status & iPD AAO (Yes: N=238, No: N=158)<sup>1</sup>** |  |  |  |
| Smoking: Yes | 4.48 | 1.01 | 1.1×10<sup>-5</sup> |
| Sex: Women | 0.18 | 1.02 | 0.861 |
| PC1 | -0.04 | 0.72 | 0.958 |
| PC2 | -0.47 | 0.70 | 0.501 |
| **Interaction between smoking status & MGS on iPD AAO (Yes: N=238, No: N=158)<sup>2</sup>** |  |  |  |
| Smoking: Yes | 4.42 | 1.00 | 1.3×10<sup>-5</sup> |
| MGS | -1.89 | 0.80 | 0.018 |
| Sex: Women | 0.05 | 1.02 | 0.964 |
| PC1 | -0.08 | 0.72 | 0.910 |
| PC2 | -0.46 | 0.69 | 0.509 |
| Smoking:MGS | 2.29 | 1.02 | 0.025 |
| **Association between MGS & iPD AAO in non-smokers (N=158)<sup>3</sup>** |  |  |  |
| MGS | -1.87 | 0.89 | 0.038 |
| Sex: Women | 2.13 | 1.77 | 0.230 |
| PC1 | -0.21 | 1.18 | 0.857 |
| PC2 | -0.34 | 1.14 | 0.766 |
| **Association between MGS & iPD AAO in smokers (N=238)<sup>3</sup>** |  |  |  |
| MGS | 0.50 | 0.58 | 0.386 |
| Sex: Women | -1.50 | 1.22 | 0.218 |
| PC1 | -0.07 | 0.90 | 0.935 |
| PC2 | -0.61 | 0.86 | 0.482 |

iPD=idiopathic Parkinson's disease, MGS=Mitochondrial polygenic score, AAO=Age at onset
R-Formulas:
<sup>1</sup>glm(formula = AAO ~ Smoking status + Sex + PC1 + PC2, family = gaussian)
<sup>2</sup>glm(formula = AAO ~ Smoking status × MGS + Sex + PC1 + PC2, family = gaussian)
<sup>3</sup>glm(formula = AAO ~ MGS + Sex + PC1 + PC2, family = gaussian)
Baseline categories: Sex=Men, Smoking status=No

Stratifying the PPP cohort by smoking status, we observed a stronger correlation between a higher MGS and earlier AAO in persons who did not smoke (r=-0.15, p=0.059; β=-1.87, SE=0.89, p=0.038) compared to persons who did smoke (r=0.053, p=0.414; β=0.50, SE=0.58, p=0.386; **Figure 2B** and **Table 4**). Longitudinally, we did not observe an association between high MGS or smoking with MDS-UPDRS Part III or MoCA, and there was also no evidence for an interaction.

## Discussion

### Genetic screening of the PPP cohort

In this study, we investigated the genetic landscape of the Dutch PPP cohort, a deeply phenotyped group of 507 persons who were followed longitudinally for 2 years. We demonstrated that PD genetics in PPP are shaped by high-impact pathogenic variants, strong risk factors in *GBA1*, and complex polygenic factors, highlighting the multifactorial etiology of PD. Approximately 18% of participants with PD in the PPP cohort carried rare variants in established PD genes. This percentage included established pathogenic variants in LRRK2 but also risk variants in *GBA1*, monoallelic *PRKN*/*PINK1* variants, as well as VUS in our thorough genetic description. Consistent with previous reports, pathogenic or likely pathogenic LRRK2 variants were the most frequent truly monogenic cause of PD, accounting for ∼1% of PPP participants with PD^3, 31^. On the other hand, we demonstrated that *GBA1* variants are the most frequent genetic finding in the PPP cohort (i.e., 15%), which is in line with previous findings^32^. The combination of targeted short-read gene panel sequencing, genome-wide genotyping, and *GBA1* long-read sequencing enabled a comprehensive and reliable genetic assessment. One advantage of combining multiple genetic assessments is that detected variants can be validated through an independent process, thereby securing the genetic finding and, if possible, avoiding labor-intensive Sanger sequencing. Furthermore, the complementary nature of long-read sequencing enabled the detection of exonic SVs in the *GBA1* gene that were missed by genotyping and short-read panel sequencing. It is known that the assessment of the *GBA1* gene is complicated by the nearby pseudogene *GBA1P*. High sequence homology to *GBA1P* can affect the mapping of short reads and lead to false-negative results^26, 33, 34^. Long-read sequencing with reads that span the entire *GBA1* gene has been shown to benefit a more comprehensive genetic *GBA1* assessment significantly in large PD cohorts ^26, 35^. Indeed, the ∼50 bp deletion in exon 9 of *GBA1* was identified only by long-read sequencing in two persons within the PPP cohort, demonstrating that clinically relevant variants remain undetected with conventional technologies.

No homozygous *PRKN, PINK1* or DJ1 variants were found, aligning with the overall late-onset phenotype of the cohort (mean AAO=56.2 years) and supporting that classic recessive early-onset PD was not represented. The slightly (non-significantly) younger average AAO among genetically positive individuals (≈1.5 years) highlights the modest phenotype shift associated with high-impact variants as previously reported^3^. The approximately one-year earlier AAO among *GBA1* variant carriers, together with the stepwise decrease in AAO across increasing variant pathogenicity, is consistent with a dose-dependent effect of *GBA1* on disease onset and aligns with previous reports linking *GBA1* pathogenic variants to earlier PD manifestation^3^. While no association with longitudinal motor severity was observed here, the trend toward greater cognitive decline in carriers of pathogenic variants further supports a known link of *GBA1* variants with cognitive decline rather than motor progression^36, 37^. Interestingly, although effect sizes were small and statistical power was limited due to the low number of variant carriers, the model estimates pointed in a similar direction for pathogenic and likely pathogenic variants compared with *GBA1* risk variants (i.e., p.Glu365Lys and p.Thr408Met). Previous studies have reported that *GBA1* risk variants are associated with a disease progression more comparable to iPD^38^. It is important to note that the longitudinal analyses in the present study were based on three assessment time points over a two-year period. Recent publications suggest that more pronounced differences in disease progression for *GBA1* variant carriers may emerge after approximately five years of follow-up^39^. Consequently, re-evaluating disease trajectories stratified by *GBA1* variant status at later time points may be critical once additional longitudinal assessments become available. Renewed follow-up assessments, again with a deep phenotyping approach, are currently being performed in all consenting survivors of the PPP cohort, at about 8 years of follow-up after the baseline assessment.

### Missense variants in *SNCA* and *CHCHD2*

In addition to demonstrating the methodological applicability of our genetic screening approach, we identified two rare missense variants in PD-associated genes, *SNCA* p.Ala124Thr and *CHCHD2* p.Ile80Val, that remain insufficiently characterized with respect to PD pathogenicity. Although both genes have been implicated in autosomal-dominant PD, we provide evidence that warrants cautious interpretation of these specific variants. Neither variant was enriched among people with PD across external datasets, and neither had robust prior literature supporting a disease association. It is important to note that *CHCHD2* p.Ile80Val was observed in both a person with PD and an elderly healthy control, while *SNCA* p.Ala124Thr was absent from large PD sequencing resources and curated databases.

In conclusion, the available genetic, population, and literature-based evidence is insufficient to support definitive pathogenic or benign classification for either variant. Nevertheless, reporting these rare variants in the genetic summary of the PPP cohort and (re)-investigation of family members is important to facilitate potential future re-evaluation should additional evidence emerge to clarify their role in PD.

### Interaction between mitochondrial polygenic score and smoking on age at onset of iPD

Even in persons without a genetic finding (iPD subgroup, N=414), complex genetic contributions remained relevant. The mitochondrial-function polygenic score (MGS) showed a small trend toward earlier AAO overall, though the effect size was minimal in the full cohort, consistent with prior publications^6, 7^. Furthermore, in independently calculated MGS specific to the oxidative phosphorylation system was associated with earlier AAO as well ^8^.

Our previous work highlighted the importance of gene-environment interactions when assessing mitochondrial polygenic burden^6^. Interestingly, particularly in non-smokers, a higher MGS was associated with earlier AAO in persons with iPD from a different PD cohort. The interaction between MGS and smoking in persons with iPD represents an independent replication of previous work from our group, strengthening the evidence that mitochondrial genetic burden interacts with lifestyle modifiers.

However, no causal conclusions can be drawn regarding whether smoking directly affects mitochondrial function in persons with PD. An alternative explanation for the observed MGS-smoking interaction is that individuals with higher MGS may enter the premotor stage of the disease earlier and, consequently, are less likely to smoke. This may be due to reduced dopamine levels, decreased sensitivity to nicotine, and the presence of an anxiety-prone pre-PD phenotype.

Consequently, more data are required to assess whether the MGS has a greater impact on non-smokers in the absence of potential neuromodulatory effects. Further studies should aim to elucidate how environmental/lifestyle modifiers may interact with mitochondria, and whether dopaminergic neuron stimulation may improve mitochondrial function in PD.

### Strengths and limitations

Although our study demonstrates a feasible and cost-effective genetic screening strategy for large PD cohorts, there are still limitations and possible causal variants that we might have missed. Our pipeline does not detect large *SNCA* multiplications, which remain challenging to identify even with long-read whole-genome sequencing^40^. In addition, homozygous or compound-heterozygous structural variants in *PRKN* or *PINK1* or rare long-range structural rearrangements spanning *GBA1*^41^ or *PRKN*^42^ are not entirely covered by our genetic assessment. Furthermore, the list of genes associated with PD is constantly expanding, with *RAB32* as a famous example for a recently discovered new PD gene^43, 44^. We used a targeted gene panel that focused only on genes known to cause PD at that time, including *GBA1*. Still, this study establishes a robust foundation of high-confidence variants for the PPP cohort, enabling downstream stratification and targeted clinical trial recruitment. Additionally, this data may help explain variability in AAO, PD symptom severity, or future disease progression.

In conclusion, our study demonstrates the applicability of combining PD-variant screening, comprehensive *GBA1* analysis, and assessment of polygenic factors to capture the genetic landscape of PD within cohorts such as the PPP cohort. These findings show that both high-impact and complex polygenic factors, as well as potential gene-lifestyle interactions, are of importance in PD, highlighting the need for integrative approaches for PD risk assessment and precision medicine.

## Data Availability

All data produced in the present study are available upon reasonable request to the authors

## Acknowledgment

The study was supported by funding from the Joint Programme of Neurodegenerative Diseases (JPND, ControlPD), by ZonMw Open (MiGut, Project number: 09120012110098) and by the Deutsche Forschungsgemeinschaft with a Heisenberg Grant (TR 1714/7-1) to JT. ZL and PM were supported by the FUN-MITO grant by the Michael J. Fox Foundation (MJFF).

