## Supplementary for "Genetic landscape of Parkinson’s disease in the Personalized Parkinson Project cohort"

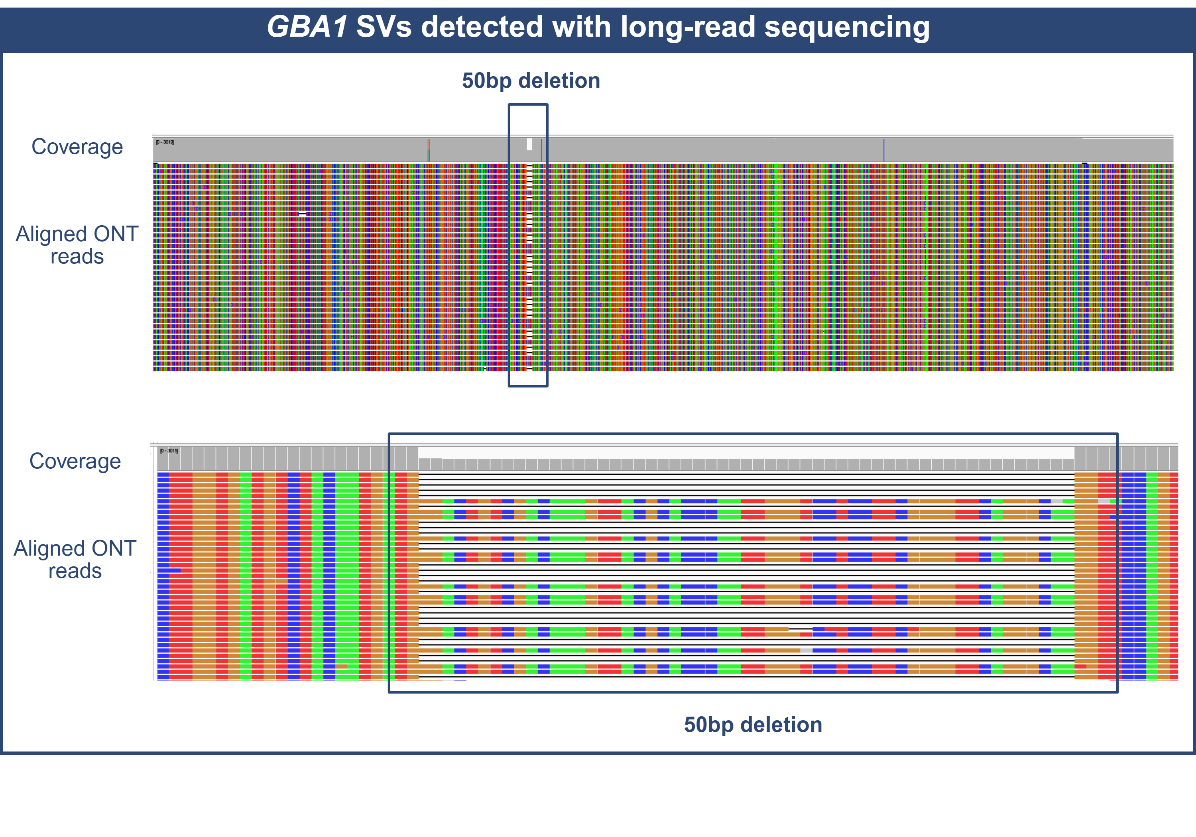

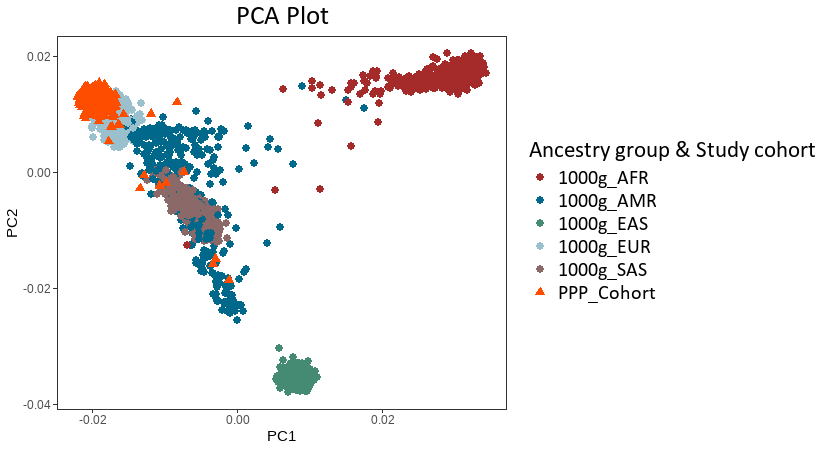
**Supplementary Material:**

**Supplementary Figure 1.** **Principal component analysis (PCA**). The PCA plot displays the clustering of samples. The PCA was derived from common SNPs that overlapped in all datasets. The samples were colored by their study cohort and their ancestry group. In addition to the PPP cohort, we used the publicly available 1000 Genomes Project dataset for validation.

1000 Genomes Project super populations: AFR=African, EAS=East Asian, AMR=Admixed American, EUR=European and SAS=South Asian

**Supplementary Figure 2. Visualization of Nanopore long-read sequencing data aligned to the GBA1 gene using the Integrative Genome Viewer (IGV).** The top panel shows the aligned long across the ~9 kb amplicon, and the lower one an enlarged ~80 bp region of the deletion. The colored bars below represent individual aligned long reads. At the exon 9 locus, a ~50 bp deletion is evident as a local reduction in coverage and as a consistent gap in multiple aligned reads, indicating the presence of a heterozygous structural variant supported by long-read sequencing.

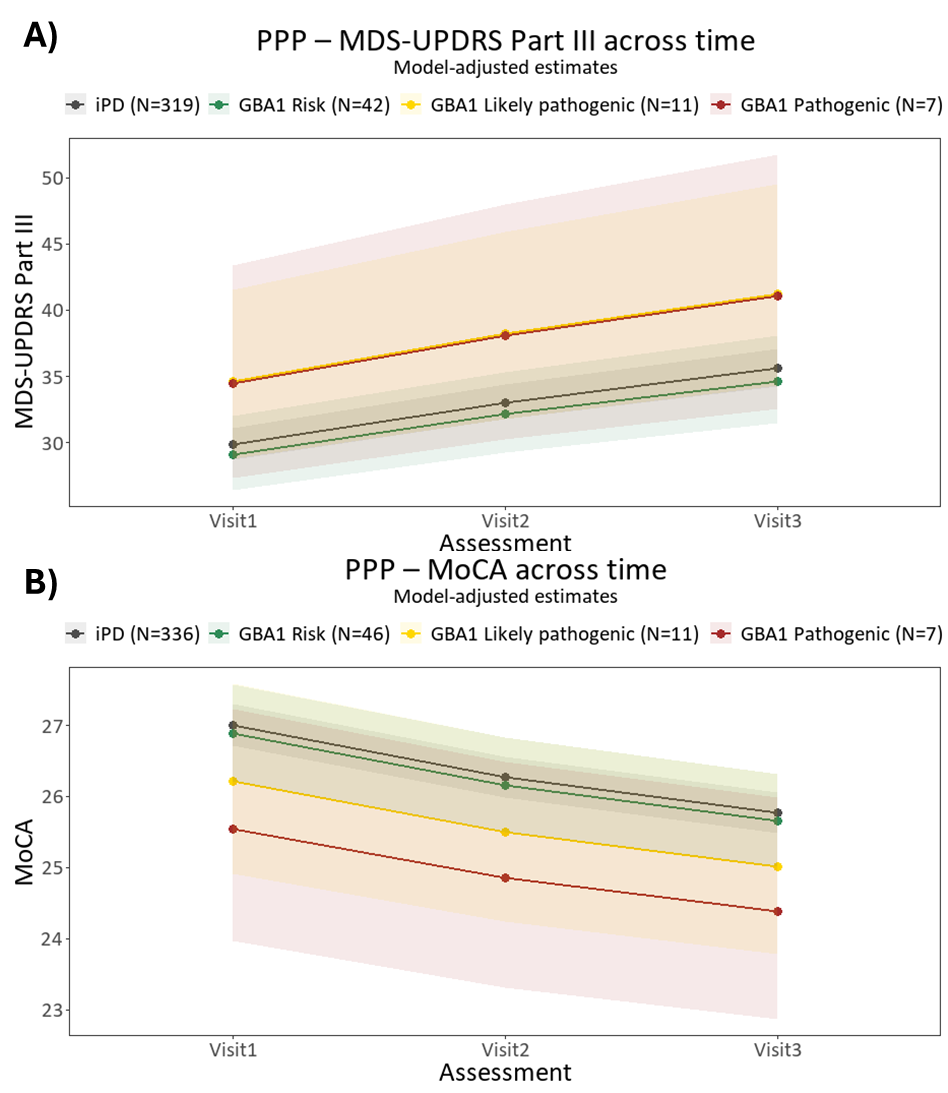

**Supplementary Figure 3. Longitudinal assessment of motor signs and cognitive performance in patients with *GBA1* variants and idiopathic PD.** Model-adjusted mean Movement Disorder Society Unified Parkinson’s Disease Rating Scale Part III (MDS-UPDRS Part III) score **(A)** or Montreal Cognitive Assessment (MoCA) scores **(B)** are shown across three longitudinal visits for patients with idiopathic PD (iPD, gray), GBA1 risk variants (green), likely pathogenic variants (yellow), and pathogenic variants (red). Shaded areas represent the 95% confidence intervals, reflecting the variability of scores over time. iPD=Idiopathic Parkinson’s disease; N=Number of individuals.

Mixed linear effect model R-formula: lmer(log(MDS-UPDRS Part III or MoCa) ~ Visit + GBA1 variant status + Sex + mfp transformed AAO + mfp transformed disease duration + z-standardized PC1 + z-standardized PC2 + (1|Patient ID))

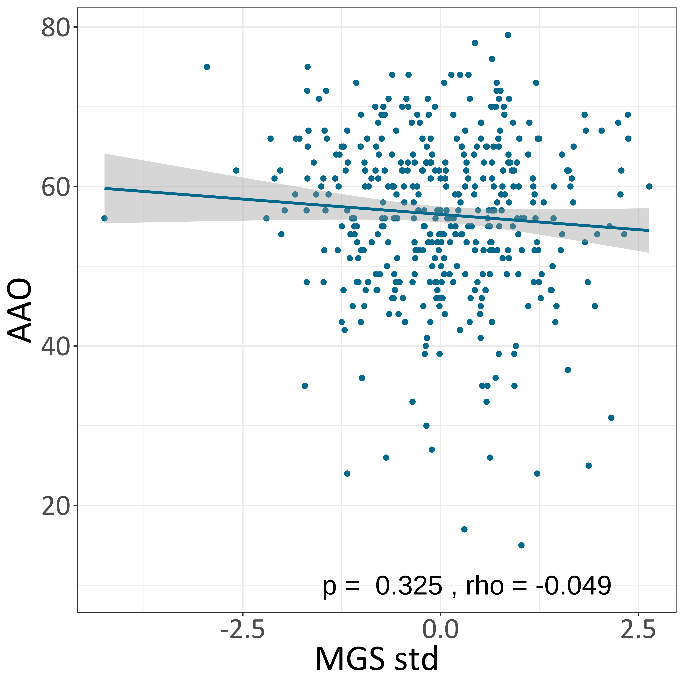

**Supplementary Figure 4. Relationship between the mitochondrial polygenic score (MGS) and age of idiopathic PD onset (AAO)**. The correlation plot shows the association between MGS and AAO in patients with idiopathic Parkinson's disease (iPD). rho = Spearman's rank correlation coefficient, p = Spearman's exploratory P-value.

**Supplementary Text 1:**

We conducted a literature review on the association between Synuclein Alpha (SNCA) and Parkinson’s disease (PD). For this purpose, we screened PubMed using the following search term:

("SNCA" OR "Synuclein Alpha" OR "Alpha-Synuclein" OR "α-Synuclein" OR "PARK1" OR "PARK4" OR "PD1" OR "NACP" OR "Non-A4 component of amyloid precursor" OR "Synuclein, Alpha (Non A4 Component Of Amyloid Precursor)" OR "Non-A beta component of AD amyloid" OR "Synuclein Alpha-140" OR "I+/--Synuclein") AND ("Parkinson" OR "Parkinson's Disease" OR "PD" OR "Parkinsonism" OR Parkinson*) AND ("SNCA Ala124" OR "SNCA A124" OR "SNCA p.Ala124Thr" OR "p.Ala124Thr" OR "A124T" OR "SNCA A124T" OR "SNCA Ala124Thr" OR "c.370G>A" OR "NM_000345.5:c.370G>A" OR "NP_000336.1:p.Ala124Thr" OR "SNCA p.Ala124Ser" OR "p.Ala124Ser" OR "A124S" OR "SNCA A124S" OR "SNCA Ala124Ser" OR "c.370G>T" OR "NM_000345.5:c.370G>T" OR "NP_000336.1:p.Ala124Ser") AND (english[Language]).

Our search identified no validated SNCA p.Ala124Thr variant. However, we found one report of p.Ala124Ser described in a Finnish cohort (<https://onlinelibrary.wiley.com/doi/pdf/10.1111/ane.13613>) in a patient with Vascular Dementia (VaD) and Schizophrenia. This variant is not currently established as pathogenic, and evidence for its role in PD remains limited.

We also screened the MDSGene database for this substitution, and no entries related to it were found (https://www.mdsgene.org/d/1/g/2?action=plot_genetic&fc=0&_mu=1&_country=1), suggesting that our study may be the first to describe this variant.

**Supplementary Text 2:**

We performed a literature review on the association of Coiled-Coil-Helix-Coiled-Coil-Helix Domain Containing 2 (CHCHD2) with Parkinson’s disease (PD). We utilized the following search term to screen PubMed:

("CHCHD2" OR "Coiled-Coil-Helix-Coiled-Coil-Helix Domain Containing 2" OR "C7orf17" OR "Chromosome 7 Open Reading Frame 17" OR "MIX17B" OR "MIX17 Homolog B" OR "MNRR1" OR "Mitochondrial Nuclear Retrograde Regulator 1" OR "Aging-Associated Gene 10 Protein" OR "PARK22" OR "NS2TP" OR "HCV NS2 Trans-Regulated Protein") AND ("Parkinson" OR "Parkinson's Disease" OR "PD" OR "Parkinsonism" OR "Parkinson*") AND (english[Language]).

The screening resulted in 118 publications. We excluded N=89 for the following reasons: reviews (N=23), pre-prints (N=2), *in vitro* studies on stem cells and iPSCs (N=4), *in vivo* studies on animal models (N=16), and publications that focused on the functional, regulative, and mechanistic aspects of CHCHD2 in both PD and other neurological diseases. Thus, we include N=29 publications. The results are summarized in **Supplementary Table 1**.

**Supplemental Table 1.** Overview of studies about the association of Coiled-Coil-Helix-Coiled-Coil-Helix Domain Containing 2 (CHCHD2) with Parkinson’s disease (PD).

| N° | Ref | Summary | Association with PD |
| --- | --- | --- | --- |
| 1 | Sun, YM., Zhou, XY., Liang, XN. *et al.* The genetic spectrum of a cohort of patients clinically diagnosed as Parkinson’s disease in mainland China. *npj Parkinsons Dis.* 9, 76 (2023). https://doi.org/10.1038/s41531-023-00518-9 | In a cohort of 832 **Chinese Han** patients clinically diagnosed with PD, including 636 early-onset (<50 years) and 196 familial late-onset (≥50 years) cases, genetic testing was performed using target sequencing of 116 movement-disorder–related genes (including 22 PD-related genes), whole-exome sequencing, multiplex ligation-dependent probe amplification (MLPA) for CNV detection, and repeat-expansion testing. 1 Heterozygous, AD Pathogenic or likely pathogenic (P/LP) variant was identified in 1 PD patient with EO: **p.Arg145Gln** (0.16%). | Yes |
| 2 | Caritativo, Erin Camille A. et al. Genetic screening of Filipinos suspected with familial Parkinson's disease: A pilot study Parkinsonism Relat Disord. 2023 Mar:108:105319. doi: 10.1016/j.parkreldis.2023.105319. Epub 2023 Feb 7. | This pilot study explored genetic mutations associated with familial PD among **Filipinos**. 18 patients from 11 families underwent genetic screening for 23 PD-related genes. The study identified PINK1 and PRKN as major contributors to early-onset familial PD in Filipinos, while **no variants** were reported in CHCHD2 or the other analyzed genes. | --- |
| 3 | Zheng Jiang et al. Production of a human iPSC line from an early-onset Parkinson’s disease patient with a novel CHCHD2 gene truncated mutation, Stem Cell Research,  Volume 64, 2022, 102881, ISSN 1873-5061, https://doi.org/10.1016/j.scr.2022.102881. | A human induced pluripotent stem cell (iPSC) line was generated from peripheral blood mononuclear cells of a Han Chinese family member with EOPD. Mutation analysis by Sanger sequencing identified a truncated CHCHD2 variant (**p.Pro53Alafs*38**), which differs from the more commonly reported missense mutations and provides a better model for studying haploinsufficiency-related mitochondrial dysfunction in PD. | Yes |
| 4 | Vacchiano, V.; Bartoletti-Stella, A.; Rizzo, G.; Avoni, P.; Parchi, P.; Salvi, F.; Liguori, R.; Capellari, S. Frequency of Parkinson’s Disease Genes and Role of *PARK2* in Amyotrophic Lateral Sclerosis: An NGS Study. *Genes* 2022, *13*, 1306. https://doi.org/10.3390/genes13081306 | A study conducted on **Italian** patients investigated the frequency of PD-related gene variants in amyotrophic lateral sclerosis (ALS) and Alzheimer’s disease (AD) compared to healthy controls (HCs) using next-generation sequencing of multiple genes, including CHCHD2. Among 130 ALS, 100 AD, and 1686 HC participants, CHCHD2 variants were identified in 1 ALS case (0.8%; c.101C>T, **p.Pro34Leu**), 1 AD case (1%; c.134C>G, **p.Ser45Cys**), and 7 HCs (0.4%). | --- |
| 5 | Yong-Ping Chen et al. The mutation spectrum of Parkinson-disease-related genes in early-onset Parkinson's disease in ethnic Chinese Eur J Neurol. 2022 Nov;29(11):3218-3228. doi: 10.1111/ene.15509. Epub 2022 Aug 4. | A study conducted on 704 early-onset PD patients (≤45 years; 110 autosomal dominant, 63 autosomal recessive, and 531 sporadic; mean age at onset 39.1 ± 6.0 years) from an ethnic **Chinese** population used whole-exome sequencing, gene dosage, and genotype–phenotype/haplotype analyses to investigate 26 PD-related genes. Two truncated variants in CHCHD2 (**p.P53Afs*38** and **p.Y99X**) were identified. They were found in 2 patients (2/110; 1.8%) with AD inheritance sharing the same haplotype. (0.20%) of CHCHD2 in Sporadic early onset PD patients was found. | Yes |
| 6 | Ping Hua et al. Genetic Analysis of Patients With Early-Onset Parkinson's Disease in Eastern China. Front Aging Neurosci. 2022 May 11:14:849462. doi: 10.3389/fnagi.2022.849462. eCollection 2022. | A study conducted on 155 unrelated PD patients (8 familial; 5 autosomal dominant, 3 autosomal recessive, and 147 sporadic; mean age at onset 43.39 ± 7.13 years) from an ethnic **Eastern Chinese** population used whole-exome sequencing and MLPA analyses to investigate 24 PD-related genes. One P/LP nonsense heterozygous mutation in CHCHD2 (**c.297C > A (p.Y99X)**) was identified in one AD PD patient (1/5 0,2%). | Yes |
| 7 | Ran Zheng ez al. Analysis of rare variants of autosomal-dominant genes in a Chinese population with sporadic Parkinson's disease. Mol Genet Genomic Med. 2020 Oct;8(10):e1449. doi: 10.1002/mgg3.1449. Epub 2020 Aug 14. | A **Chinese** study of 191 sporadic PD patients (106 early-onset, 85 late-onset; mean age at onset 51.15 ± 11.16 years) and 200 ethnicity-matched controls analyzed 12 autosomal-dominant PD genes through panel sequencing. A CHCHD2 pathogenic variant (**p.Thr61Ile**) and a candidate variant (**p.Pro2Leu**) were identified in 2 patients (2/191; 0,01%) and 1 control (1/200; 0,005% ). Regarding CHCHD2 p.Thr61Ile, the carrier of this mutation also carried the LRRK2 p.Ala419Val mutation. | Yes |
| 8 | Sumeet Kumar, et al. Novel and reported variants in Parkinson's disease genes confer high disease burden among Indians. Parkinsonism Relat Disord. 2020 Sep:78:46-52. doi: 10.1016/j.parkreldis.2020.07.014. Epub 2020 Jul 16. | A comprehensive study of 250 PD from **India**, including 206 sporadic and 44 familial cases (227 early-onset, 23 late-onset), employed whole-exome sequencing to examine 20 PD-associated genes. The cohort was evenly recruited from South India (n = 124) and North India (n = 126), providing a representative population sample. Within the autosomal-dominant PD genes, a heterozygous CHCHD2 variant (**p.G73V**) was identified in a single patient (1/250, 0.4%), with an age at onset of between 30 and 35 years. | --- |
| 9 | Nannan Li, Ling Wang, Jinhong Zhang, Eng-King Tan, Junying Li, Jiaxin Peng, Liren Duan, Chaolan Chen, Dong Zhou, Li He, Rong Peng,  Whole-exome sequencing in early-onset Parkinson's disease among ethnic Chinese, Neurobiology of Aging, Volume 90, 2020, Pages 150.e5-150.e11, ISSN 0197-4580, https://doi.org/10.1016/j.neurobiolaging.2019.12.023. | A total of 240 **Chinese** PD patients were included in the study, comprising 193 sporadic PD cases and 47 familial EOPD cases (39 ADPD and 7 ARPD). Whole-exome sequencing was performed to screen PD genes including CHCHD2, one likely pathogenic variant (**p.T61I**, heterozygous) was identified in one autosomal-dominant PD patient **(1/39)**. | Yes |
| 10 | Nannan Yang et al.  Systematically analyzing rare variants of autosomal-dominant genes for sporadic Parkinson's disease in a Chinese cohort, Neurobiology of Aging, Volume 76, 2019, Pages 215.e1-215.e7, ISSN 0197-4580, https://doi.org/10.1016/j.neurobiolaging.2018.11.012. | A **Chinese** study was conducted on 1456 sporadic PD patients, including 736 early-onset (<50 years) and 720 late-onset (≥50 years) cases. A total of 1568 unrelated age- and sex-matched healthy controls were included in the study. Targeted sequencing of seven AD-PD genes (SNCA, LRRK2, GIGYF2, VPS35, EIF4G1, DNAJC13, and CHCHD2) was performed using MIP capture. 72 rare nonsynonymous-coding variants with minor allele frequency ≤0.1% were identified. Among them, 2 **CHCHD2** variants were detected; one of uncertain significance and one likely pathogenic: **p.A79S**, which was absent in controls. These variants were found in 2/1456 cases and 1/1568 controls. | Yes |
| 11 | Danielle D. Voigt et al, CHCHD2 mutational screening in Brazilian patients with familial Parkinson's disease, Neurobiology of Aging, Volume 74, 2019, Pages 236.e7-236.e8, ISSN 0197-4580, https://doi.org/10.1016/j.neurobiolaging.2018.09.026. | This study conducted the first molecular analysis of the CHCHD2 gene in a cohort of 122 index cases from **Brazilian** families with autosomal dominant forms of PD. Genomic DNA was isolated from peripheral blood and the 4 exons of the *CHCHD2* gene, and their intron-exon boundaries were analyzed by bidirectional Sanger sequencing. Knowing that mutations in *SNCA*, *LRRK2*, *VPS35*, and *GBA1* genes were previously excluded in these patients, **no pathogenic or risk variants were found**, suggesting that genetic variants of *CHCHD2* are not a common cause of familial PD in Brazilian patients. | --- |
| 12 | Richard G. Lee et al, Early-onset Parkinson disease caused by a mutation in CHCHD2 and mitochondrial dysfunction. Neurol. Genet. 4, e276 (2018), https://doi.org/10.1212/NXG.0000000000000276 | A **Caucasian** woman in her 30’s presented with EOPD with no significant family history. Genetic analysis using whole-exome sequencing and/or a targeted neuromuscular panel, followed by confirmatory bidirectional Sanger sequencing, identified a novel homozygous missense mutation in **CHCHD2 (c.211G>C; p.Ala71Pro)**. Her unaffected parents were both heterozygous carriers of this variant, while her healthy sister did not carry the mutation. | Yes |
| 13 | Aya Ikeda et al, A novel mutation of CHCHD2 p.R8H in a sporadic case of Parkinson's disease, Parkinsonism & Related Disorders, Volume 34, 2017, Pages 66-68, ISSN 1353-8020, https://doi.org/10.1016/j.parkreldis.2016.10.018. | In a cohort of 4 PD patients from Sado Island in **northern Japan**—three with autosomal dominant PD and one sporadic case with an age of onset in their 30’s. Using Sanger sequencing and MLPA to screen SNCA multiplications, LRRK2 (exons 11, 21, 31, 41), parkin variants, and CHCHD2 missense mutations, the researchers identified a novel **CHCHD2** variant **(c.23G>A; p.R8H)** exclusively in the sporadic PD patient (1/4), with no pathogenic mutations detected in the three ADPD cases. | Yes |
| 14 | Elisa Rubino et al,  Genetic analysis of CHCHD2 and CHCHD10 in Italian patients with Parkinson's disease, Neurobiology of Aging, Volume 53, 2017, Pages 193.e7-193.e8, ISSN 0197-4580, https://doi.org/10.1016/j.neurobiolaging.2016.12.027. | In an **Italian** cohort of 119 PD patients, including both familial and sporadic cases. All coding regions and flanking splice sites of CHCHD2 were PCR-amplified and sequenced by Sanger analysis after excluding individuals with known SCNA, LRRK2, *PINK1*, or *PARK7* mutations. **No** previously reported rare CHCHD2 mutations were detected (0/119), although one sporadic late-onset PD patient carried the intronic CHCHD2 variant rs199529573 (c.300+31G>A). | --- |
| 15 | Monica Gagliardi et al, Analysis of CHCHD2 gene in familial Parkinson's disease from Calabria, Neurobiology of Aging, Volume 50, 2017, Pages 169.e5-169.e6, ISSN 0197-4580, https://doi.org/10.1016/j.neurobiolaging.2016.10.022. | In 165 familial PD cases from **Southern Italy**, CHCHD2 genetic screening—after excluding other PD-related gene mutations—revealed only intronic variants (rs816408, rs10043, rs816407, rs8406) occurring at similar frequencies in patients and controls. **No** pathogenic CHCHD2 mutations were identified (0/165). | --- |
| 16 | Cristina Tejera-Parrado et al,  Genetic analysis of CHCHD2 in a southern Spanish population, Neurobiology of Aging, Volume 50, 2017, Pages 169.e1-169.e2, ISSN 0197-4580, https://doi.org/10.1016/j.neurobiolaging.2016.10.019. | A total of 536 PD cases from **Southern Spain** (76 familial PD and 101 early-onset PD) were screened for CHCHD2 variants using high-resolution melting, followed by Sanger sequencing, genotyping, and case-control association analysis. **No** pathogenic CHCHD2 mutations were detected (0/536). Only four non-exonic variants were identified: three in the 5′UTR (c.–9T>G [rs10043], c.–34C>A [rs816407], c.–65T>G [rs816406]) and one in the 3′UTR (c.*125G>A [rs8406]). | --- |
| 17 | Chao Gao et al, Mutation analysis of CHCHD2 gene in Chinese Han familial essential tremor patients and familial Parkinson's disease patients,  Neurobiology of Aging, Volume 49, 2017, Pages 218.e9-218.e11, ISSN 0197-4580, https://doi.org/10.1016/j.neurobiolaging.2016.10.001. | A study of 133 **Chinese Han** patients with familial autosomal dominant PD and 211 healthy controls sequenced all four exons and exon–intron boundaries of CHCHD2, identifying 5 variants: 4 non-coding (c.-11G>A, c.-9T>G, c.456+125G>A, c.456+179T>C) and one coding variant (**rs142444896 5C>T, p.Pro2Leu**). **No pathogenic variants were detected**. A smaller analysis in 171 individuals with familial essential tremor identified only non-coding or splice-region variants and no coding mutations. | --- |
| 18 | Li N-N, Wang L, Tan E-K, Cheng L, Sun X-Y, Lu Z-J, Li J-Y, Zhang J-H, Peng R. 2016. Genetic Analysis of CHCHD2 Gene in Chinese Parkinson's Disease. Am J Med Genet Part B 171B: 1148–1152. https://doi.org/10.1002/ajmg.b.32498 | A study of 1,058 **Chinese** PD patients (31 familial, 1,027 sporadic, and 1,095 controls analyzed CHCHD2 variants using PCR-LDR genotyping confirmed by direct sequencing. All subjects were homozygous for rs10043, and **1,052/1,058** were **homozygous** for the **p.Pro2Leu** variant. Only **9/1,058** subjects (6 PD and 3 controls) were **heterozygous** for **p.Pro2Leu**. This study shows a twofold increase of p.Pro2Leu in PD, suggesting it may be a **risk factor in Asian populations**. | Yes (Risk factor) |
| 19 | Hongwei Wu et al, Genetic analysis of the CHCHD2 gene in a cohort of Chinese patients with Parkinson disease, Neuroscience Letters, Volume 629, 2016, Pages 116-118, ISSN 0304-3940, https://doi.org/10.1016/j.neulet.2016.06.054. | In a cohort of 162 **Chinese** PD patients (90 familial ADPD and 72 sporadic PD), all coding exons of the CHCHD2 gene were amplified by PCR and sequenced. 4 non-coding variants (c.-34C>A, c.-9T>G, c.*125G>A), including 1 novel variant, c.*154A>G, were identified. This last was detected in a heterozygous state in 2/90 familial ADPD patients and absent in controls. One coding variant, **c.5C>T (Pro2Leu)**, was found in a single sporadic PD patient. **No pathogenic** CHCHD2 mutations were observed in this cohort **(0/162).** | --- |
| 20 | Xinglong Yang et al, Mutational scanning of the CHCHD2 gene in Han Chinese patients with Parkinson’s disease and meta-analysis of the literature, Parkinsonism & Related Disorders, Volume 29, 2016, Pages 42-46, ISSN 1353-8020, https://doi.org/10.1016/j.parkreldis.2016.05.032. | A cohort of 584 **Han Chinese** PD patients (30 familial AD, 554 sporadic) and 594 controls was screened for CHCHD2 by sequencing all coding regions, exon–intron boundaries, UTRs, and flanking regions. 3 exonic missense variants were detected; **p.Pro2Leu** (2 controls/594 and 4 Sporadic PD patients/584), **p.Arg18Gln** (1 sporadic PD/584), and **p.Arg145Gln** (1 sporadic PD/584). No variant co-segregated with PD in families; **No** pathogenic CHCHD2 mutations in this cohort. 12 variants in untranslated/flanking regions were also identified. | Yes |
| 21 | Qian Lu et al, Mutation analysis of the CHCHD2 gene in Chinese Han patients with Parkinson's disease, Parkinsonism & Related Disorders, Volume 29, 2016, Pages 143-144, ISSN 1353 8020, https://doi.org/10.1016/j.parkreldis.2016.04.012. | A total of 245 **Chinese Han** PD patients (110 familial, 135 sporadic) and 220 controls from mainland China were screened for CHCHD2 using PCR and direct sequencing covering all coding regions and exon–intron boundaries. Three known non-coding variants: c.–34C>A (rs816407), c.–11G>A (rs200226056), and c.–9T>G (rs10043) were detected in both patients and controls, and **no coding mutations** were identified. | --- |
| 22 | Eva Koschmidder, Anne Weissbach, Norbert Brüggemann, Meike Kasten, Christine Klein, Katja Lohmann, Neurology. 2016 Feb 9;86(6):577-9. doi: 10.1212/WNL.0000000000002361. Epub 2016 Jan 13. | A cohort of 330 PD patients (mean AAO 54.7 ± 18.1 years; 47% with a positive family history), predominantly of **German** ancestry (90%), underwent Sanger sequencing of all four CHCHD2 exons and exon–intron boundaries. A likely pathogenic nonsense variant in exon 3 (**c.376C>T; p.Gln126X**), predicted to produce a truncated protein, was identified in one German patient (male, AAO in his 40’s) : **(1/330)**. Two frequent non-coding variants, rs10043 (c.-9T>G) and rs8406 (c.*125G>A), were also detected. | Yes |
| 23 | Tian-Sin Fan, Hang-I. Lin, Chin-Hsien Lin, Ruey-Meei Wu, Lack of CHCHD2 mutations in Parkinson's disease in a Taiwanese population, Neurobiology of Aging, Volume 38, 2016, Pages 218.e1-218.e2, ISSN 0197-4580, https://doi.org/10.1016/j.neurobiolaging.2015.11.020. | A total of 723 **Taiwanese** PD patients, including 137 familial cases (86 autosomal dominant, 51 autosomal recessive) and 586 sporadic cases, along with 710 controls, were screened using MLPA for major PD genes and mutations of Parkin, PINK1, DJ-1, ATP13A2, PLA2G6, FBXO7, LRRK2, and HtrA2 were excluded. Sequencing of all 4 exons and exon-intron boundaries of CHCHD2 was performed. 5 CHCHD2 variants were identified: 3 non-coding (3′UTR +125G>A [rs8406], and 2 novel variants +179T>C and +300A>G) and 2 coding variants; p.G83G (0 familial PD/137; 1 control/129) and the known p.Pro2Leu (2 familial PD/137; 2 controls/129). **No** pathogenic CHCHD2 mutations were found **(0/137).** | --- |
| 24 | Chang-he Shi et al, CHCHD2 gene mutations in familial and sporadic Parkinson's disease, Neurobiology of Aging, Volume 38, 2016,  Pages 217.e9-217.e13, ISSN 0197-4580, https://doi.org/10.1016/j.neurobiolaging.2015.10.040. | In a **Chinese** ADPD family comprising 7 affected members, investigators first performed whole-exome sequencing in 2 patients and 1 control, followed by sequencing of the full CHCHD2 coding region and exon–intron boundaries. After excluding 18 known PD-related gene mutations, they identified a heterozygous CHCHD2 variant (c.182C>T; p.Thr61Ile) segregating with disease: present in (2/2) PD patients and 1 essential tremor case, absent in unaffected relatives and in 500 controls.  To further assess the contribution of CHCHD2 in their population, they next sequenced all CHCHD2 exons in an additional 382 PD patients (18 ADPD, 364 sporadic) and 384 controls. **No** additional pathogenic mutations were detected **(0/382)**; only five known SNVs were found across UTR, intronic, and flanking regions, including **Pro2Leu.** No other potentially disease-causing mutations were found in familial and sporadic PD patients. | Yes |
| 25 | Ming Zhang et al, Mutation analysis of CHCHD2 in Canadian patients with familial Parkinson's disease, Neurobiology of Aging, Volume 38, 2016, Pages 217.e7-217.e8, ISSN 0197-4580, https://doi.org/10.1016/j.neurobiolaging.2015.10.038. | A cohort of 155 **Canadian** familial PD patients underwent Sanger sequencing of all CHCHD2 exons. Only one rare 5′UTR variant (c.–12C>T; rs112876794) was detected in a single patient, and three coding CHCHD2 polymorphisms **(Pro2Leu, Pro14Ser, Ile118Met)** were homozygous for the major allele in an additional 85 screened PD patients. **No** pathogenic CHCHD2 mutations were identified (0/155). All cases had been pre-screened to exclude mutations in PARK2, DJ-1, SNCA, PINK1, LRRK2, VPS35, and GBA, and previously reported CHCHD2 mutations from Japanese and Caucasian cohorts were absent in this Canadian familial PD group. | --- |
| 26 | Ogaki K et al, Mitochondrial targeting sequence variants of the CHCHD2 gene are a risk for Lewy body disorders. Neurology. 2015 Dec 8;85(23):2016-25. doi: 10.1212/WNL.0000000000002170. Epub 2015 Nov 11. PMID: 26561290; PMCID: PMC4676755. | Across three cohorts—878 **US Caucasian** PD patients (356 familial), 335 **Irish** PD patients (16 familial), and 394 **Polish** PD patients (58 familial)—together with their respective 717, 365, and 350 controls, all CHCHD2 coding exons and exon–intron boundaries were sequenced. A total of 23 CHCHD2 variants were identified across the combined datasets; including 9 exonic variants; 4 located in exon 1 (p.P2L, p.G4R, p.P14S, p.A16A) and 5 in exon 2 (p.V31V, p.P34L, p.A37V, p.A49V, and p.A93V) and none in exons 3 and 4. 5 5'UTR variants, 6 variants in the 3′UTR and 3 variants in intron 3 were also identified, but **no** definitively pathogenic CHCHD2 mutations were found in the US (0/878), Irish (0/355), or Polish (0/394) PD series. | --- |
| 27 | Zhenhua Liu et al, Mutation analysis of CHCHD2 gene in Chinese familial Parkinson's disease, Neurobiology of Aging, Volume 36, Issue 11, 2015, Pages 3117.e7 3117.e8, ISSN 0197-4580, https://doi.org/10.1016/j.neurobiolaging.2015.08.010. | Genetic analysis of mutations in the *CHCHD2* gene was conducted in a cohort of 92 families with autosomal dominant PD from **mainland China**. **No** mutations in *CHCHD2* gene were identified (0/92), suggesting that CHCHD2 mutations might not be a common cause of PD in Chinese familial cases. | --- |
| 28 | Iris E Jansen et al, CHCHD2 and Parkinson's disease, The Lancet Neurology, Volume 14, Issue 7, 2015, Pages 678 679, ISSN 1474-4422, https://doi.org/10.1016/S1474-4422(15)00094-0. | Across 1,243 PD cases and 6,927 controls of **European ancestry** from the International Parkinson's Disease Genomics Consortium (IPDGC), exome sequencing revealed none of the previously reported Asian-specific CHCHD2 variants (Thr61Ile, Arg145Gln, and 300+5G>A), supporting their rarity outside Asian populations. Instead, three novel exon-2 variants (**Ala32Thr, Pro34Leu, and Ile80Val**) were identified in **4/1,243 PD** cases (two French and two American), all with familial and early-onset PD, and were absent in controls. Additional non-coding variation included two 5′UTR and two 3′UTR variants. NeuroX array analysis also detected the Pro2Leu (rs142444896) variant, previously reported in Japanese cohorts. Overall, the lack of exonic variants in controls suggests that CHCHD2 variation may represent a **rare risk factor** for PD in individuals of **Western European ancestry**. | Yes (Risk Factor) |
| 29 | Manabu Funayama et al, CHCHD2 mutations in autosomal dominant late-onset Parkinson's disease: a genome-wide linkage and sequencing study, The Lancet Neurology, Volume 14, Issue 3, 2015, Pages 274-282,  ISSN 1474-4422, https://doi.org/10.1016/S1474-4422(14)70266-2. | In a **Japanese** cohort of 341 ADPD index cases obtained from the DNA bank of the Comprehensive Genetic Study on Parkinson’s Disease and Related Disorders, exome sequencing and whole-genome sequencing, followed by Sanger validation, identified 3 CHCHD2 mutations in 4/341 independent families. These included two missense mutations—182C>T **(Thr61Ile)** detected in 2/341 cases and 434G>A **(Arg145Gln)** detected in 1/341—and one splice-site mutation **(300+5G>A)** identified in 1/341 cases. None of these variants were found in controls. | Yes |

**Supplementary Table 2.** Demographic overview of patients with PD who carried a rare variant in genes associated with autosomal-dominant PD.

|  | **AAE range**  **[years]** | **AAO range**  **[years]** | **Disease duration** | **Sex** | **Ancestry** |
| --- | --- | --- | --- | --- | --- |
| **LRRK2 p.Gly2019Ser** | | | | | |
| **1.** | 50-59 | 50-59 | 2 years | Woman | European |
| **2.** | 60-69 | 60-69 | 3 years | Woman | European |
| **3.** | 70-79 | 70-79 | 7 years | Woman | European |
| **SNCA p.Ala124Thr** | | | | | |
| **1.** | 50-59 | 40-49 | 8 years | Woman | European |
| **CHCHD2 p.Ile80Val** | | | | | |
| **1.** | 60-69 | 60-69 | 5 years | Woman | European |

N=Number of participants, AAO=age at onset of initial motor symptoms, AAE= age of initial examination

**Supplementary Table 3.** The association between *GBA1* variant status and motor signs or cognitive impairment over time in the PPP cohort. The Movement Disorder Society Unified Parkinson’s Disease Rating Scale Part III (MDS-UPDRS Part III) or the Montreal Cognitive Assessment (MoCA) scores were evaluated longitudinally over time and assessed with a mixed linear model in patients with *GBA1*-PD and patients with iPD.

|  | **Estimate** | **SE** | **p-value** |
| --- | --- | --- | --- |
| **Association of *GBA1* variant status and MDS-UPDRD Part III (iPD: N=319, *GBA1* Risk: N=42, *GBA1* Likely pathogenic: N=11, *GBA1* Pathogenic: N=7)** | | | |
| **Visit 2** | 0.10 | 0.02 | 2.4×10^-9^ |
| **Visit 3** | 0.18 | 0.02 | <2.0×10^-16^ |
| ***GBA1* Risk** | -0.03 | 0.05 | 0.586 |
| ***GBA1* Likely pathogenic** | 0.15 | 0.09 | 0.125 |
| ***GBA1* Pathogenic** | 0.14 | 0.12 | 0.232 |
| **Sex: Women** | -0.18 | 0.03 | 3.5×10^-8^ |
| **Disease duration** | -0.36 | 0.05 | 7.5×10^-14^ |
| **AAO** | 0.51 | 0.17 | 0.003 |
| **PC1 std** | 0.001 | 0.02 | 0.966 |
| **PC2 std** | -0.01 | 0.02 | 0.645 |
| **Association of *GBA1* variant status and MoCA (iPD: N=336, *GBA1* Risk: N=46, *GBA1* Likely pathogenic: N=11, *GBA1* Pathogenic: N=7)** | | | |
| **Visit 2** | -0.03 | 0.01 | 3.1×10^-7^ |
| **Visit 3** | -0.05 | 0.01 | <2.0×10^-16^ |
| ***GBA1* Risk** | -0.004 | 0.01 | 0.746 |
| ***GBA1* Likely pathogenic** | -0.03 | 0.03 | 0.255 |
| ***GBA1* Pathogenic** | -0.06 | 0.03 | 0.090 |
| **Sex: Women** | 0.04 | 0.01 | 8.2×10^-7^ |
| **Disease duration** | 0.02 | 0.01 | 0.109 |
| **AAO** | -0.26 | 0.05 | 1.3×10^-8^ |
| **PC1 std** | 2.0×10^-6^ | 0.01 | 1.00 |
| **PC2 std** | 1.9×10^-4^ | 0.01 | 0.975 |

*N*=Number of individuals, Formula in R (package lme4): lmer(log(MDS-UPDRS Part III or MoCa) ~ Visit + GBA1 variant status + Sex + mfp transformed AAO + mfp transformed disease duration + z-standardized PC1 + z-standardized PC2 + (1|Patient ID))

Baseline categories: Visit=Visit 1 (i.e., assessment at enrolment), *GBA1* variant status: No variant/idiopathic PD, Sex: Men
